# Electronic health record signatures identify undiagnosed patients with Common Variable Immunodeficiency Disease

**DOI:** 10.1101/2022.08.03.22278352

**Authors:** Ruth Johnson, Alexis V. Stephens, Sergey Knyazev, Lisa A. Kohn, Malika K. Freund, Leroy Bondhus, Brian L. Hill, Tommer Schwarz, Noah Zaitlen, Valerie A. Arboleda, Manish J. Butte, Bogdan Pasaniuc

**Author notes:** These authors contributed jointly (Manish J. Butte, Bogdan Pasaniuc).

## Abstract

One of the most common human inborn errors of immunity (IEI) is Common Variable Immunodeficiency (CVID), a heterogeneous group of disorders characterized by a state of functional and/or quantitative antibody deficiency and impaired B-cell responses. Although over 30 genes have been associated with the CVID phenotype, over half the CVID patients have no identified monogenic variant. There are currently no existing laboratory or genetic tests to definitively diagnose CVID and none are expected to be available in the near future. The extensive heterogeneity of CVID phenotypes causes patients with CVID to face a 5 to 15 years of delay in diagnosis and initiation of treatment, leading to a critical diagnosis odyssey. In this work, we present PheNet, an algorithm that identifies patients with CVID from their electronic health record data (EHR). PheNet computes the likelihood of a patient having CVID by learning phenotypic patterns, *EHR-signatures*, from a high-quality, clinically curated list of bona fide CVID patients identified from the UCLA Health system (N=197). The prediction model attains superior accuracy versus state-of-the-art methods, where we find that 57% of cases could be detected within the top 10% of individuals ranked by the algorithm compared to 37% identified by previous phenotype risk scores. In a retrospective analysis, we show that 64% of CVID patients at UCLA Health could have been identified by PheNet more than 8 months earlier than they had been clinically diagnosed. We validate our approach using a discovery dataset of ∼880K patients in the UCLA Health system to identify 74 of the top 100 patients ranked by PheNet score (top 0.01% PheNet percentile) as highly probable to have CVID in a clinical blinded chart review by an immune specialist.

## Introduction

Human inborn errors of immunity (IEI), also referred to as primary immunodeficiencies (PIDs), are rare, often monogenic diseases that confer susceptibility to infection, autoimmunity, and auto-inflammation [1]. There are currently over 400 distinct IEIs and dozens more are discovered each year due to the availability of whole exome or genome sequencing[1]. One of the most common IEIs is the Common Variable Immunodeficiency (CVID) phenotype, a heterogeneous group of disorders characterized by a state of functional and/or quantitative antibody deficiency and impaired B cell responses[2,3]. The most common clinical presentation of CVID includes recurrent sinopulmonary infections, but can also include a variety of symptoms related to autoimmunity (e.g., autoimmune hemolytic anemia) and immune dysregulation (e.g., enteritis, granulomata). The prevalence of the CVID phenotype ranges from 1 in 10,000 to 1 in 50,000 individuals worldwide[4]. Only ∼2,000 known cases in the United States have been identified as of 2019[5], suggesting that between 5,000 to 33,000 patients with CVID have yet to be found in the United States alone.

Despite advances in genome sequencing technologies and the increased capacity of diagnosis for IEIs, the spectrum of genetic etiologies of the CVID phenotype is not fully understood. Over 30 genes have been implicated in CVID, but the specific genetic cause is not identified for the majority of individuals. More recently, it has even been proposed that the genetic basis of CVID can be described by a polygenic genetic architecture, where cumulative genetic effects across the genome confer disease risk[6,7]. Because there is no clear causal genetic mechanism, there is no genetic test available for providing definitive diagnoses. Furthermore, this genetic variability leads to heterogeneous presentations of patients with CVID, making it even more difficult to diagnose. Individuals with CVID present with broad phenotypic patterns of autoimmunity and/or infection susceptibility. Since the immune system is intertwined with nearly all organs and tissues, the clinical presentation of rare immune diseases such as CVID intersects with virtually every medical specialty. This causes the fragmentation of patients across multiple clinical subspecialties, which leads to significant delays in diagnosis and treatment. This consequential delay is one of the major challenges in initiating clinical care for CVID patients, averaging 5 years in children [8] to 15 years in adults [9]. This protracted delay in treatment increases both morbidity and mortality [9–11]. Thus, there is a highly critical and unmet need to reduce the diagnostic delay for CVID and promptly provide these patients with treatments such as immunoglobulins and immunomodulators.

The recent availability of large-scale electronic health records (EHR) has enabled the computational assessment of patients’ phenotypic characteristics solely based on their medical records[12–16], enabling the systematic and scalable review for millions of individuals. A fundamental difficulty in this approach is having *a priori* knowledge about how the patterns of CVID are represented solely through EHR. We refer to these patterns describing the manifestations of CVID through EHR as the *EHR-signatures* of the disease. Because there is not a single clinical presentation for CVID, constructing an EHR-signature for CVID is not straightforward. In this work, we present a computational algorithm, PheNet, that computes a CVID score to prioritize patients likely to have CVID and thus candidates for immune specialist review and formal diagnoses. We leverage a high-quality, clinically curated list of CVID patients (N=197) identified within the UCLA Health system to construct a statistical model to learn the EHR-signature of CVID. Given the low prevalence of CVID and the complexities associated with diagnosing patients, this curated dataset represents one of the largest databases of CVID patients, enabling us to construct models previously not available due to the limited sample sizes of rare disease patients.

We demonstrate that PheNet attains superior accuracy versus state-of-the-art methods. We find that 57% of cases could be detected within the top 10% of all individuals ranked by PheNet in the EHR whereas previous phenotype risk scores specific to CVID [12] only capture 37% of cases, and prior genetic testing risk scores[13] only capture 23% of cases. Using EHR data from UCLA Health medical records, we show in a retrospective analysis that 64% of previously undiagnosed CVID patients at UCLA could have been identified by PheNet over 8 months before they received their initial diagnosis. We further validate our approach with a blinded clinical review from a clinical immunologist for the top 100 patients identified by PheNet out of a total of 880K individuals in the UCLA Health population. We find that 74% of individuals could be confirmed as highly probable as having CVID and specifically 8% of the top 100 could be confirmed as putatively diagnosed with CVID based on an examination of their full medical record. Taken together, EHR-based algorithms such as PheNet will expedite the diagnosis of CVID patients and will help identify novel phenotypic patterns of CVID.

## Results

### Summary and description of CVID cohort at UCLA

Central to our approach was training and validating our model using a dataset of individuals with a known diagnosis of CVID. To first identify a smaller set of medical records to review, we restricted the search to individuals with ICD-10 code D80.* (Immunodeficiency with predominantly antibody defects) which produced approximately N=3,200 individuals within the UCLA Health system. Medical records for each individual were then manually reviewed by a clinical immunologist to determine the significance and recurring patterns associated with the specific diagnosis codes (see Methods). This process helped eliminate individuals who received an immunodeficiency code not directly associated with CVID, for example based on acute occurrences or individuals with a cancer diagnosis who were receiving immunosuppressive medications that caused immune dysregulation. This procedure identified a cohort of N=197 individuals with a confirmed CVID diagnosis (Supplementary Figure S1). For model training and validation, we constructed a matched control cohort based on self-identified sex, self-identified race/ethnicity, age (closest within a 5-year window), and the number of days recorded in the EHR (closest within a 180-day window) (see Methods), resulting in a total of 197 cases and 1,106 controls (Figure 1).

**Figure 1:**
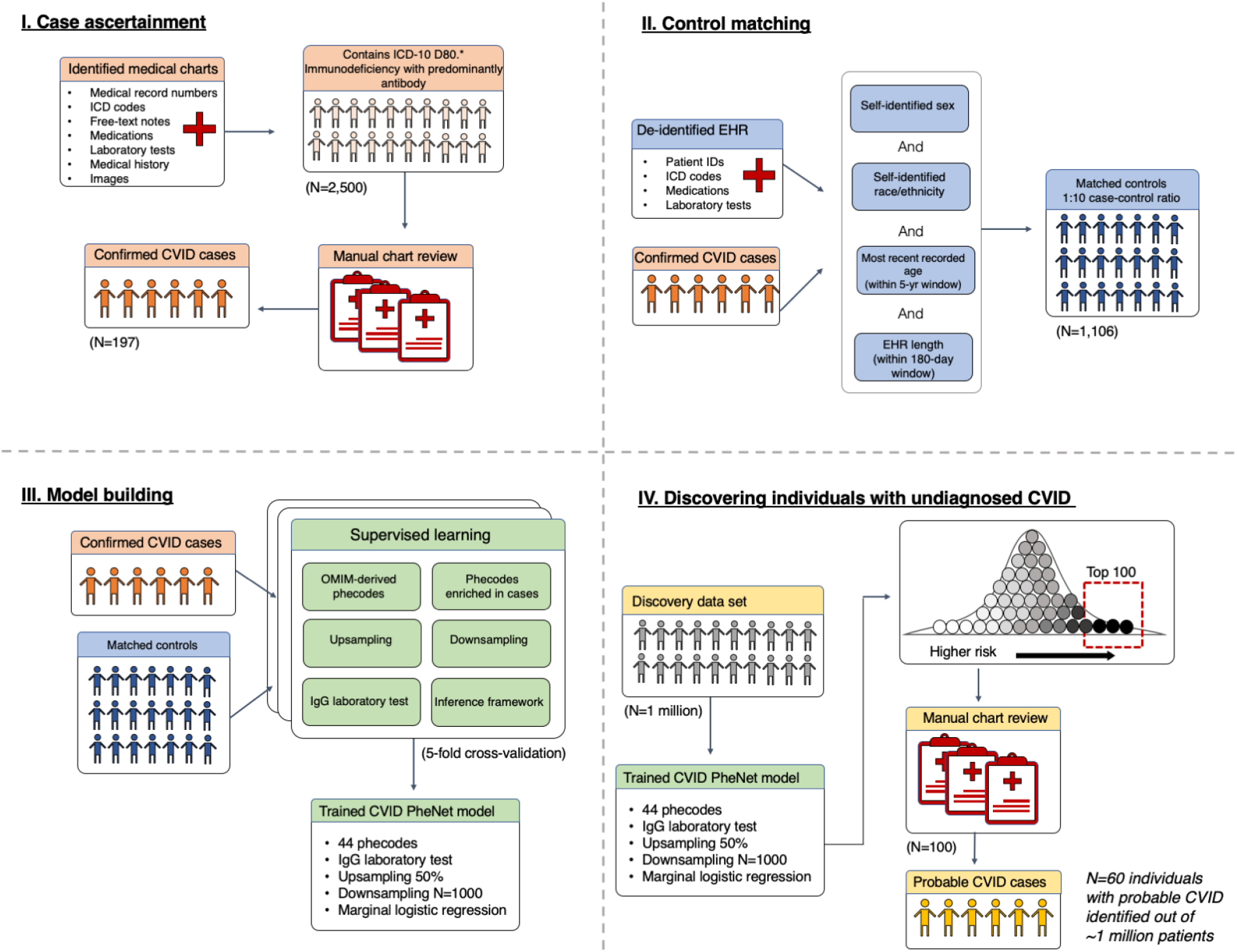
Overview of PheNet model training and application within a discovery cohort. We present a visual summary of case/control cohort construction, PheNet model training, and application within a discovery dataset at UCLA Health. In (I) we show the workflow for constructing a case cohort of clinically diagnosed patients with CVID from medical charts (N=197). (II) shows the criteria used to create a matched control cohort from the EHR (N=1,106). (III) visually summarizes the construction of a prediction model, including feature selection from phecodes, the inclusion of laboratory values, a variety of inference frameworks, and data balancing techniques. Finally, (IV) demonstrates how the PheNet model can be applied within a discovery cohort to identify patients with a high likelihood of CVID who can then be further assessed by manual chart review to confirm diagnosis.

The resulting CVID cohort was 71.6% female, and the average age was 55.4 (SD: 19.5). Previously constructed cohorts had a very similar demographic profile that also showed an increased female predominance[17]. Using the ICD-based diagnosis dates (see Methods), we found that the average age of individuals at diagnosis was 55.0 (SD: 19.0) years old which is consistent with a large portion of individuals being diagnosed with CVID after age 40 [10]. To evaluate the extent of patients’ immune dysregulation, we assess patients’ immunoglobulin G (IgG) levels, a common antibody that is typically low in CVID patients [4]. Within the case-cohort, we found that 86.3% of individuals had at least one IgG laboratory test and 37.1% had at least one abnormally low IgG laboratory test result (<600 mg/dL). Out of the control cohort, we found that only 3.2% of individuals had an IgG test and only 0.45% with an abnormal result which is consistent with the majority of individuals not likely having any immunodeficiencies. Many individuals had extensive medical history at UCLA Health where the average extent of EHR data was 14.7 years (SD: 7.9), and the average number of unique ICD-10 codes per individual was 95.8 (SD: 95.7) (Table 1, Supplementary Figure S2). However, there were 6 individuals out of the 197 CVID cases who had fewer than 10 encounters within the EHR from UCLA. This could reflect individuals who came to UCLA only to receive a formal diagnosis or those who only came for a second opinion, but largely had their care at a different medical center.

**Table 1:** Demographics of CVID case/control cohorts. We show a summary of the individuals in both the CVID case (N=197) and control cohorts (N=1,106) including age, sex, number of unique ICD codes, number of years recorded in the EHR, and immunoglobulin G (IgG) laboratory tests. If patients had more than one IgG test, the lowest value was used.

|  | **Case** | **Control** |
| --- | --- | --- |
| **Sample size** | 197 | 1,106 |
| **Age (years)** |  |  |
| Mean (s.d.) | 55.35 (19.53) | 54.20 (20.26) |
| Median | 60.00 | 57.00 |
| **Sex (%)** |  |  |
| Male | 28.43 | 30.81 |
| Female | 71.57 | 69.19 |
| **Mean number of unique ICD codes** | 95.76 (95.67) | 69.00 (95.67) |
| **Mean medical record length years** | 14.72 (7.93) | 14.27 (7.89) |
| **IgG laboratory test** |  |  |
| No test | 12.7% | 96.8% |
| IgG >600 mg/dL (normal) | 49.2% | 2.7% |
| IgG <600 mg/dL (abnormal) | 37.1% | 0.45% |

### Constructing a CVID risk score model from EHR-derived phenotypes

We used the curated set of cases to learn the EHR-signature for CVID as follows. For features, we used *phecodes[18]* (codes derived from ICD codes used to represent clinically meaningful phenotypes from the EHR) and laboratory measurements of immunoglobulin G (IgG) (see Methods). To prevent overfitting, we selected a subset of phecodes (out of possible ∼1,800 codes) that best captured the phenotypic patterns of CVID. We first selected phecodes matching the clinical description of CVID listed in the Online Mendelian Inheritance in Man (OMIM)[19] database for a total of 34 phecodes. Then, leveraging the annotated data specifically for this study, we included the top K significant phecodes with a significantly higher frequency in the cases as compared to the controls (see Methods). To prevent biases, we excluded the actual phecode for CVID itself (279.11) from the set of features.. Varying the value of K controlled the tradeoff between adding more information to the model and overfitting due to the increased data dimensionality. We found that K=10 provided a high level of performance while also preventing overfitting, and we used this parameter in all subsequent analyses (Supplementary Figure S3A, S4A). In addition to the set of selected phecodes, we included the laboratory test for immunoglobulin G (IgG) levels as that gives a proxy for the immune state of the patient. Because we are only interested in capturing whether patients have had a test and then if this result was abnormally high, we discretized laboratory measurements as a categorical variable where patients’ lab result was either normal (IgG >600 mg/dL), abnormal (IgG <600 mg/dL), or no IgG test was recorded in their medical record.

We next compared a variety of prediction methods to learn a function that best mapped the feature set to each individual’s CVID status. We evaluated various methods that varied in model complexity, including linear methods such as marginal logistic regression of each feature, penalized joint models like ridge regression, as well as non-linear methods such as random forest regression (Supplementary Figure S3B, S4B). We found that marginal regression and ridge regression achieved similarly high performance (AUC-ROC/PR(marginal): 0.95/0.83, AUC-ROC/PR(ridge): 0.96/0.88). We opted to use marginal regression to maintain the most straightforward interpretability of the regression coefficients. We additionally assessed the performance of the model when including IgG information and found that the inclusion of this single feature added a substantial increase in performance (AUC-ROC/PR(IgG): 0.95/0.83, AUC-ROC/PR(no-IgG): 0.89/0.73) (Supplementary Figure S3C, S4C). To account for the severe case imbalance associated with predicting rare diseases, we performed random upsampling of the cases to achieve a more balanced dataset. Comparing various upsampled ratios, we found that a ratio of 0.50 provided optimal performance (Supplementary Figure S3D, S4D). Our final prediction model included the 34 phecodes selected from OMIM, the K=10 phecodes learned from the case-cohort, and the IgG laboratory test with an upsampling ratio of 0.50. Using 5-fold cross-validation, we showed that the average PheNet scores for individuals with CVID had a significantly higher risk score than the matched controls (Cochran-Armitage Test test: *p*-value < 2.2×10^−16^) (Supplementary Figure S4). We emphasize that this risk score does not quantify the risk of an individual developing CVID in the future, but instead assesses whether or not the patient likely already has CVID at the present time (but has just not yet been diagnosed).

### PheNet is more accurate than existing phenotype risk scores for predicting CVID

Next we compared PheNet against PheRS, an unsupervised risk score designed for identifying undiagnosed patients with rare diseases [12], and the chromosomal microarray (CMA) risk score, a method designed to predict patients who would benefit from CMA tests for diagnoses[13]. The model for the CMA risk score was pre-computed from the Vanderbilt EHR since re-training the model would require constructing a training dataset of individuals with validated CMA tests which we did not have at this time. However, because PheRS is an entirely unsupervised method, we were able to re-train the model for CVID prediction within the UCLA EHR. We found that PheNet performed 17% better than PheRS when comparing AUC-ROC and 42% better when comparing AUC-PR. In comparison to the CMA test, PheNet performed 30% and 66% better in terms of AUC-ROC and AUC-PR (Figure 2A, 2B). In practice, only individuals with very high risk scores would be candidates for follow-up. Setting a threshold score of 0.90, we found that 57% of cases could be detected within the top 10% of individuals ranked by PheNet score (Figure 2C). In contrast, PheRS and the CMA-score only captured 37% and 23% of cases at the same threshold.

**Figure 2:**
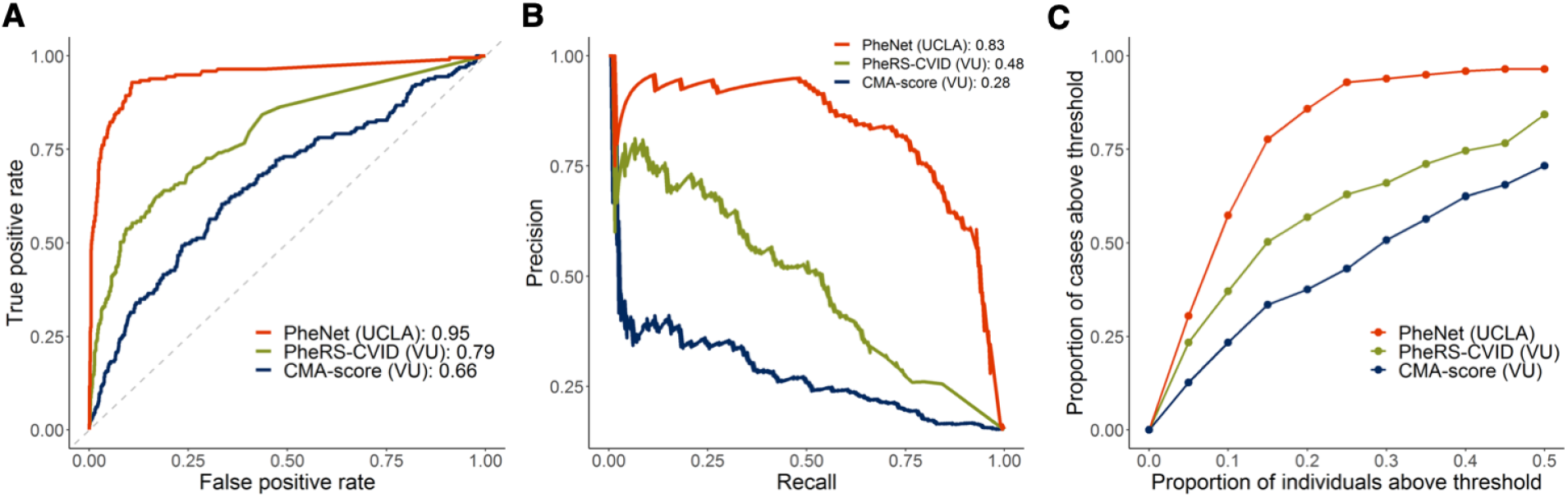
PheNet is more accurate than existing phenotype risk scores for predicting CVID. Performance metrics comparing the performance of PheNet, PheRS-CVID, and the CMA-score within the case (N=197) and control (N=1,106) cohorts from the UCLA Health population. The CMA-score was computed using weights pre-trained from data from Vanderbilt (VU); PheNet and PheRS-CVID were trained using weights trained from EHR data at UCLA. (A) and (B) show the receiver operating characteristic and precision-recall curves across the different prediction models. AUC is provided in parentheses in the legend. In (C), we display a curve showing individuals with a PheNet score > 0.90 and the proportion of CVID cases captured within the varying percentiles of PheNet scores.

The improvement of PheNet over the CMA-score is most likely because the CMA-score was developed for a highly-related but broader problem such as the identification of individuals that could benefit from CMA testing for diagnosis. The genetic cause for CVID is not found for over half of the patients, suggesting that even performing a CMA test would not likely identify CVID patients in the majority of cases. Additionally, deviations in performance could also be due to the fact that the CMA model was trained in EHRs from a separate institution. However, the CMA model was previously tested in out-of-hospital populations and showed little decrease in performance. To further investigate the potential bias due to EHR from different institutions, we compared the accuracy of PheRS using a model that was trained in the Vanderbilt EHR. Because the PheRS method is entirely unsupervised, we could do a systematic performance comparison when using models trained at UCLA and Vanderbilt (VU). We found that the pre-trained feature weight for the phecode 561.1 (Diarrhea) was not available in the Vanderbilt EHR, thus we excluded this feature from both models for this set of analyses. We found that the AUC-ROC and AUC-PR were almost exactly the same (PheRS-UCLA: 0.79, 0.48; PheRS-VU: 0.79, 0.49), suggesting that the EHR-signature for CVID is very similar between the institutions and not likely a major source of bias (Supplementary Figure S5).

We performed additional analyses to assess whether the performance of PheNet was artificially biased because scores were computed for individuals based on EHR information obtained both before and after their diagnoses. To test whether a more temporally restricted set of EHR data could still have appropriate predictive power, we created a “censored” testing dataset that limited each individual to only information present in the medical record prior to an individual’s “ICD-based diagnosis” for CVID. Because we do not have access to the exact date of patients’ formal diagnoses (due to a date shift in the EHR not present in the manually reviewed medical records), we estimate the date based on the occurrences of the ICD-10 code for CVID (D83.9) within the EHR. We clarify that the cohort of CVID patients was formally identified through manual chart review and that this ICD-based procedure was only used to identify the approximate date of diagnosis within the EHR (see Methods). The training dataset was still trained on all data points up to the present regardless of the diagnosis date as this does not affect test performance. Using this more restricted test set, we found a modest reduction in performance, but we were still able to capture a large percentage of CVID patients. Specifically, we found 46% of cases compared to 57% of cases within the top 10% of patients ranked by PheNet (Supplementary Figure S6). When comparing AUC-ROC and AUC-PR, we see a 17.7% and 51.7% decrease in performance. This drop in performance could be because some patients do not have substantial medical history at UCLA preceding their diagnosis which would drastically limit the prediction power. When limiting our assessment to CVID patients with at least 1 year of EHR data before their diagnosis (N=58), we find only an 8.1% and 44.6% drop in performance for AUC-ROC and AUC-PR respectively, suggesting that with adequate medical history, there is limited performance bias when using all EHR information up to the present.

### PheNet identifies new patients with CVID in real data

We next sought to quantify the utility of PheNet as a predictive tool for identifying whether patients could be diagnosed earlier by conducting an analysis using the UCLA EHR data from over 880K individuals at UCLA. The dataset comprised all individuals within UCLA Health with at least one encounter and at least one ICD code, for a total of 880K individuals (as of 2019), including our previously established case-cohort of 197 CVID cases. Using 5-fold cross-validation, we divided the data into 80% training set and 20% testing set folds and ran PheNet on each fold of the data (see Methods). To mirror how PheNet would be used in practice, we limited the testing dataset to only features that appeared in the EHR prior to an individual’s ICD-based diagnosis. For different scoring thresholds, we captured CVID patients at various times both before and after their diagnoses (Supplementary Figure S7). In practice, the score threshold could be chosen according to a specific goal and the amount of resources available. For example, one would recommend using a high stringency score threshold, thus capturing fewer individuals, if patients were to be followed-up individually which is a resource-expensive undertaking.

To ensure individuals had an adequate amount of medical history prior to their diagnosis of CVID, we restricted the analysis to individuals with at least one year of EHR data prior to their ICD-based diagnosis (N=58). We set a threshold PheNet score of 0.9 for this analysis and found that PheNet identified 64% of individuals with CVID before their ICD-based diagnosis (Figure 3). The median number of days between the date identified by PheNet and the date of diagnosis was 244 days (SD: 374). For example, the individual shown in Figure 4 reached the top percentile of the PheNet score distribution 41 days before their record reflected any immunodeficiency diagnosis codes. This patient had accumulated 7 phecodes that influenced their PheNet score in the years prior to diagnosis. Then the patient’s record revealed measurement of a modestly low IgG level, which further increased their risk score. This example demonstrates the advantage of aggregating information from both phenotypes and laboratory tests to identify individuals as high risk. These results show that PheNet has substantial utility for not only identifying undiagnosed individuals with CVID but also bringing them to a diagnosis earlier than they might have in a usual clinical scenario.

**Figure 3:**
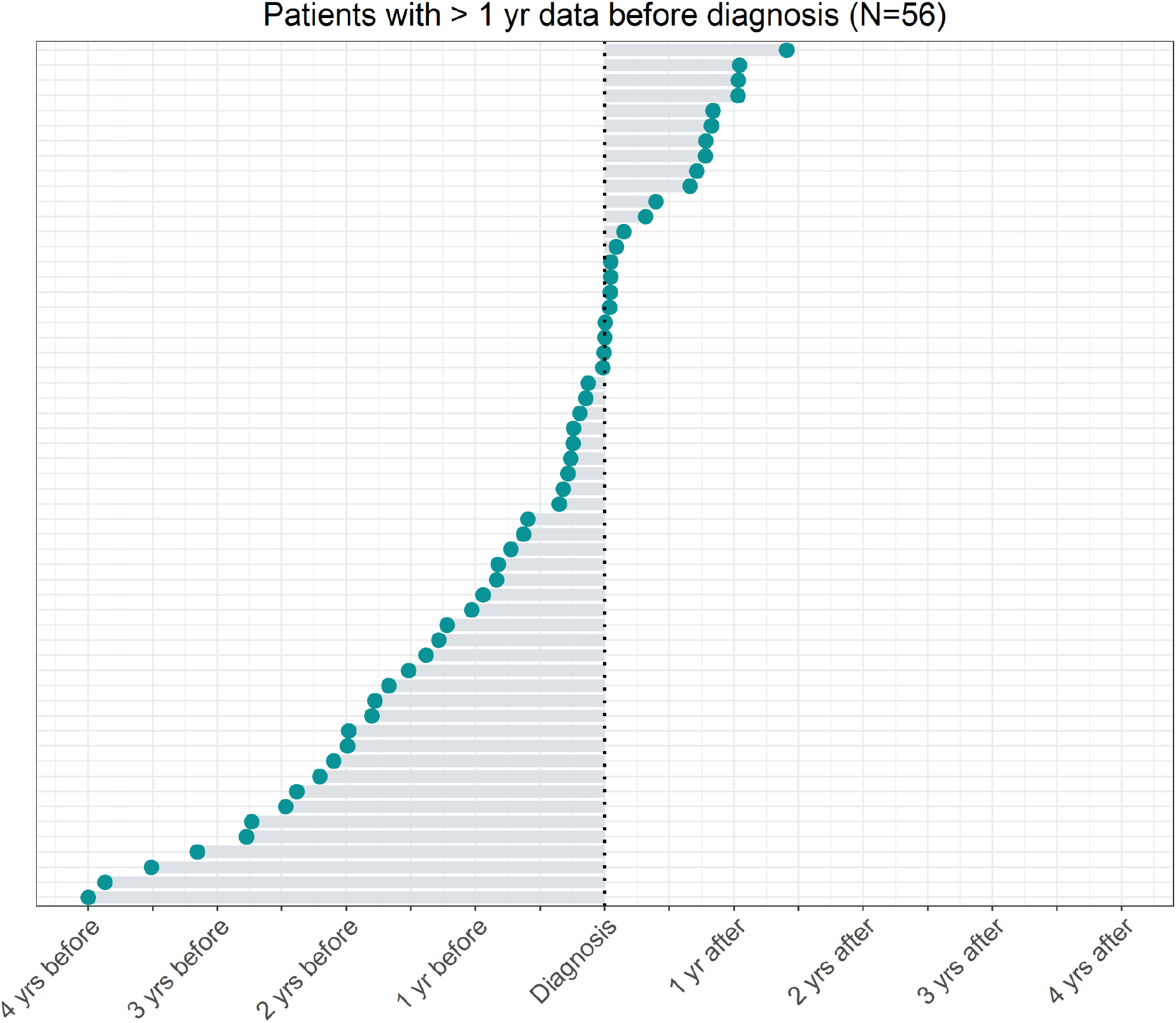
PheNet identifies CVID patients before their original diagnosis dates. Distribution of the time between individuals’ ICD-based diagnoses for CVID and the time point at which individuals’ risk score > 0.90 (denoted at the blue circles). Only individuals with at least 1 year of EHR data prior to their ICD-based diagnosis were included. Two individuals were excluded from the graph because they did not meet the score threshold at any point in time for a total of N=56 individuals shown. ICD-based diagnoses were determined as the time point when individuals first accumulate at least two CVID ICD codes (D83.9) within a year.

**Figure 3:**
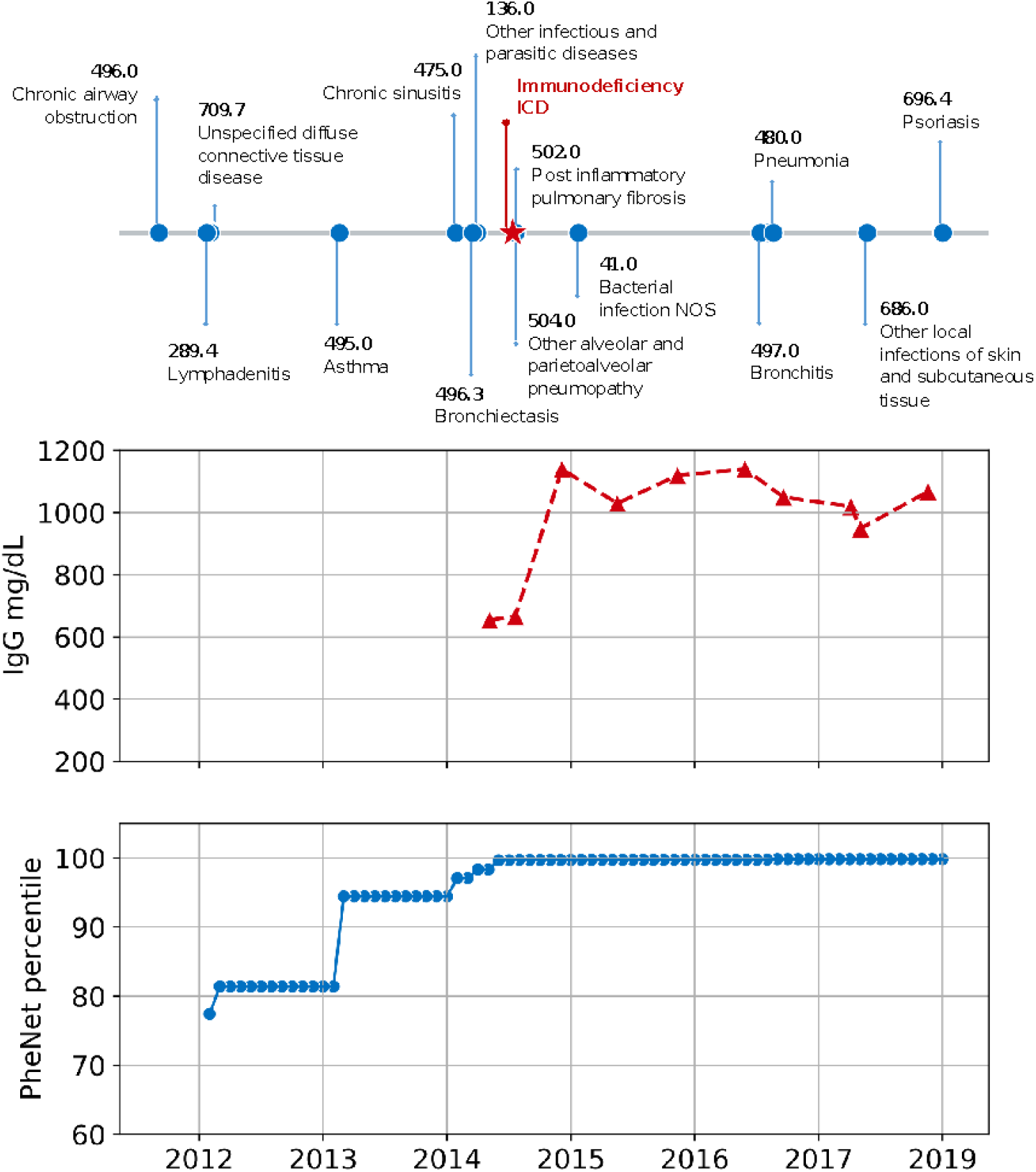
Sample patient’s CVID timeline. The top panel lists all CVID-relevant phecodes on a sample patient’s record. The point when the patient received their first immunodeficiency billing code is denoted by the red star. The middle panel shows the patient’s immunoglobulin G (IgG) laboratory results over time, where a value < 600 mg/dL is considered abnormal. The bottom panel shows the percentile of the patient’s risk score computed over time. Specifically, we show that the patient reached the 99th percentile of the PheNet score distribution 41 days before their medical record showed evidence of specific immunodeficiency care. Note that the patient’s timeline has been date-shifted.

### Clinical validation of identified undiagnosed individuals with CVID

To validate the utility of PheNet for identifying new patients with CVID, we conducted an analysis using the UCLA EHR data from over 880K individuals at UCLA as the discovery cohort. We removed from consideration all individuals who were deceased or who had phecodes corresponding to solid organ transplants, cystic fibrosis, or infection with the human immunodeficiency virus, resulting in the removal of 42,346 individuals. Individuals with these disorders may exhibit similar clinical profiles as those with CVID, but their phenotypes are likely due to their immunocompromised conditions, not a primary genetic disease. We then selected the top 100 individuals identified by PheNet and a control group of 100 randomly selected individuals from the patient population. On average, the group of top 100 individuals had an average of 15.5 years of medical history and the randomly selected group had 7.1 years (Supplementary Table S1).

We scrambled these two sets of patients and performed a clinical chart review for these individuals (see Methods). Medical records were directly examined by a clinical immunologist who was blinded to the groups and not informed that they were validating a risk score algorithm for CVID. The clinician had access to each individual’s full medical record including notes, images, and scanned documents, which were not available to the PheNet algorithm. Each individual was ranked according to an ordinal scale from 1 to 5 quantifying the likeliness of having CVID where 1 was defined as “near certainty not CVID” and 5 was “definitive as CVID” meaning that the patient meets the criteria of a physician diagnosis. From the list of top 100 ranked individuals, 74% of individuals were assigned a score of 3, 4, or 5, indicating that they were highly probable as having CVID (Figure 5). Specifically, 8% of individuals were assigned a score of 5, meaning that they were positively diagnosed with CVID in having low immunoglobulin levels and poor humoral responses to vaccine antigens or having a prior outside physician diagnosis of CVID. In contrast, the individuals who were randomly chosen exclusively had scores of 1, 2, or 3, and 90% of individuals had a score of 1 or 2, indicating that they likely did not have CVID. Overall, these results validate that our approach is useful to identify patients with CVID and overcome the major challenge of initiating care in a timely manner. The reduction of delays in diagnosis will enable patients to seek appropriate medical care to reduce morbidity and mortality.

**Figure 5:**
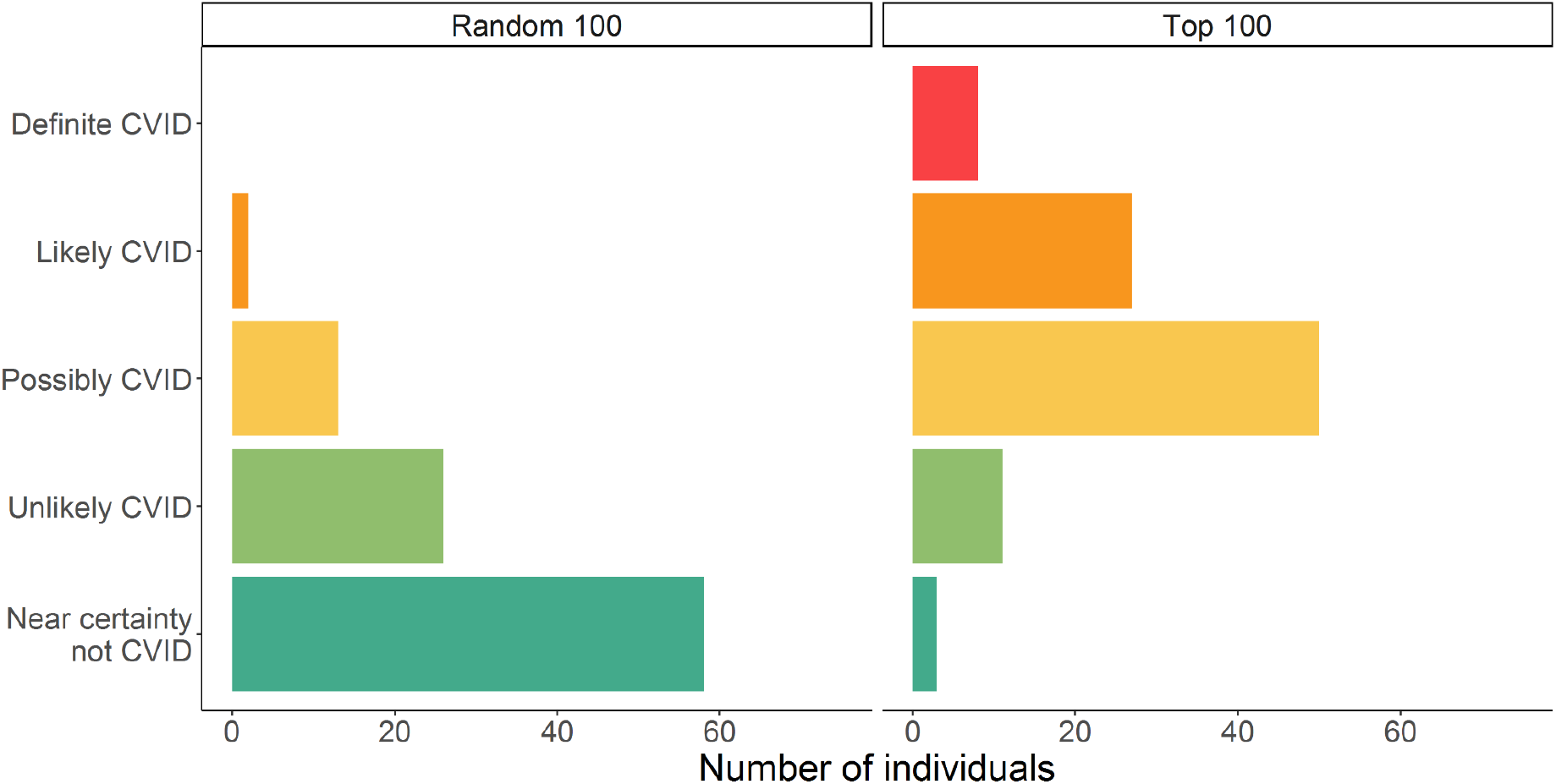
PheNet identifies undiagnosed individuals with CVID. We show the CVID clinical validation scores for the top 100 individuals with the highest PheNet score and 100 randomly sampled individuals. Each individual was ranked according to an ordinal scale from 1 to 5 quantifying the likeliness of having CVID where 1 was defined as “near certainty not CVID” and 5 was “definitive as CVID”.

In addition to prediction performance, it is also important to understand the symptoms that contributed to each individual’s increased risk status. In practice, it would not be sufficient to only identify individuals to refer to an immunology clinic, but it is also necessary to explain exactly which factors contributed to their identification. Examining the regression coefficients from the model in the form of odds ratios, we can identify phenotypes that were most predictive (Supplementary Table S2). We find that some of the most predictive features (e.g. primary thrombocytopenia) were not provided from the OMIM clinical description but were from the set of enriched phecodes identified from the case-cohort, further emphasizing the benefit of including a well-curated case-cohort in the prediction model. The signs and symptoms that contributed to each of the top 100 individuals’ risk scores are shown in Figure 6. Overall, there are wide variations in the symptoms of each individual, demonstrating the utility of methods that aggregate both numerous symptoms and laboratory results to identify patients at risk. There is no single feature present in all 100 individuals with the highest PheNet scores, underscoring the lack of any single clinical manifestation as being pathognomonic of CVID. We also observed that the majority of individuals had a mixture of both autoimmune and infection-related phecodes, further demonstrating the heterogeneity of the CVID phenotype. These patterns were consistent with those observed in the cohort of formally diagnosed CVID patients (Supplementary Figure S8). In contrast, the majority of randomly selected individuals did not have any major symptoms matching the patterns of CVID estimated by PheNet, and the signs and symptoms present within this group were those that were among the most common in the general population such as upper respiratory infections and asthma (Supplementary Figure S9).

**Figure 6:**
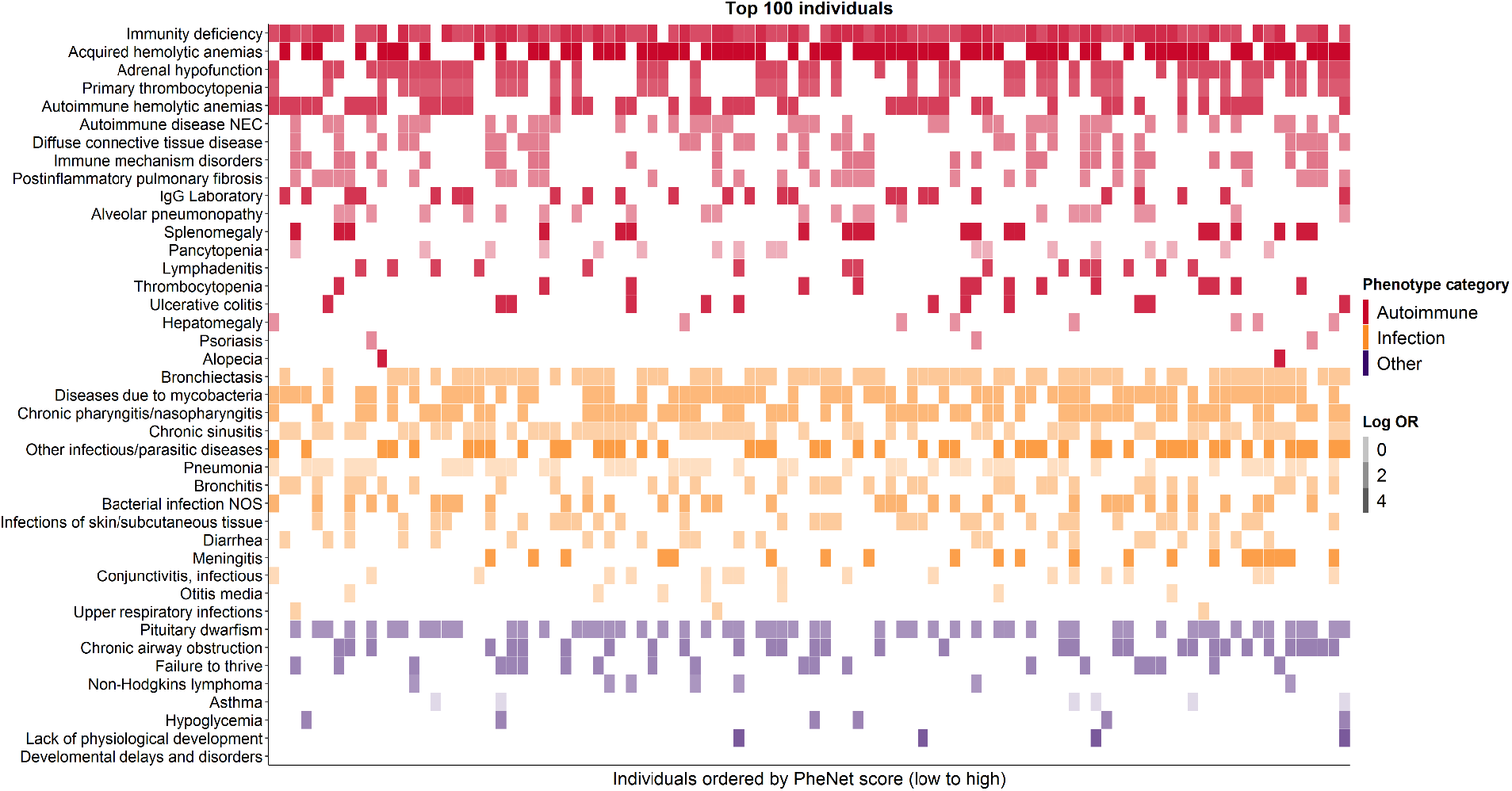
EHR-signatures of high-risk individuals for CVID. Each row shows a clinical feature from the PheNet model and each column is a patient’s EHR-profile. The top 100 individuals identified by PheNet are shown where the lowest to highest risk scores are displayed left to right. Boxes are colored according to phenotype category (autoimmune, infection, neither) and shaded according to the weight of each feature in the algorithm in the form of log odds ratio.

### Validation on the University of California wide data warehouse

For general applicability, we next tested whether PheNet could be applied to new databases. We validated the generalizability of PheNet using de-identified clinical data collected from the University of California medical centers that include (a) University of California Los Angeles; (b) University of California San Francisco; (c) University of California Davis; (d) University of California San Diego (but not Rady Children’s Hospital); and (e) University of California Irvine (> 4.9 million patient records, https://www.ucbraid.org/cords, Table 2). We scored and ranked each individual in the UC-wide data set using the PheNet weights calculated from UCLA data as above (i.e., no training was performed on the UC-wide data). To assess the utility of the scores, we asked whether PheNet could identify patients who had at least one encounter with a diagnosis code of CVID (ICD code ‘D83.9’) (N=1,838 out of >4.9M patient records). When ranking patients by PheNet scores, we found a striking enrichment of patients with a diagnosis code of CVID in the top-ranked patients. For example, among the top 10,000 patients ranked by PheNet score, we found 44% for UCLA to 64% for UCD of all patients with a CVID diagnosis code among more than 2 million patient records. A random ranking of patients that would find less than 6 patients from each clinical site in the top 10,000 patients. This result demonstrates an enrichment of CVID cases among those with high PheNet scores and showcases the power of this approach to prioritize patients suspected for CVID for follow-up analyses. Taken together, these results confirmed that PheNet maintains robust interoperability with new databases and new data formats after training PheNet with UCLA’s CVID patients.

**Table 2:** Enrichment of patients with a CVID diagnosis code identified within the UC-Wide Data Warehouse. The number of patients with the CVID ICD-10 code (D83.9) in each top PheNet scored cohort across the UC-Wide Data Warehouse.

|  | UCLA | UCSF | UCD | UCSD | UCI |
| --- | --- | --- | --- | --- | --- |
| Total patients | 1,642,284 | 1,222,352 | 794,585 | 733,128 | 579,360 |
| Total patients with CVID ICD code | 1,018 | 580 | 355 | 98 | 372 |
| PheNet top 100 | 10<br>(1.72%) | 10<br>(2.82%) | 6<br>(6.12%) | 15<br>(4.03%) | 17<br>(3.93%) |
| PheNet top 1,000 | 63<br>(10.9%) | 57<br>(16.1%) | 32<br>(32.7%) | 53<br>(14.3%) | 80<br>(18.5%) |
| PheNet top 5,000 | 178<br>(30.7%) | 131<br>(36.9%) | 50<br>(51.0%) | 139<br>(37.4%) | 206<br>(47.6%) |
| PheNet top 10,000 | 255<br>(44.0%) | 184<br>(51.8%) | 63<br>(64.3%) | 188<br>(50.5%) | 272<br>(62.8%) |

## Discussion

In this work, we identified phenotypic patterns of CVID, or EHR-signatures, encoded in patients’ medical records and trained an algorithm to identify patients who likely have CVID but who have been otherwise “hiding” in the medical system. Due to the heterogeneity of clinical presentations for IEI phenotypes, CVID patients can initially present to a wide range of clinical specialists who focus on the specific organ system involved (e.g., the lung) rather than directly to an immunologist for the underlying immune defect. This organ-based approach of our current healthcare system can result in tunnel vision and hinder a formal diagnosis in IEI, particularly for those patients who have multi-system manifestations that fluctuate over time. As a result, these patients face a diagnostic delay of 10 or more years. Each year of delay in the diagnosis of CVID results in an increase in infections, antibiotic use, emergency room visits, hospitalizations, and missed days of school and work totaling over USD $108,000 compared to the year after the diagnosis of CVID is made (in 2011 dollars, which is roughly $145,000 in 2022 dollars) [20]. The diagnostic delay for adults with CVID ranges between 10-15 years[21], suggesting that $1M or more per CVID patient is being misdirected by the US healthcare system because of diagnostic delays. Considering that there may be ∼10,000 or more individuals currently waiting to be diagnosed with CVID, the aggregate impact to the US health system is billions of dollars due to a failure to diagnose CVID in a timely fashion. Beyond the economic impact, the non-quantifiable impacts on patients’ lives due to diagnostic delays are even more significant. For example, previous studies have shown that undiagnosed patients suffer from anxiety and depression as they undergo costly tests and specialty visits. [20]

Prompt identification of patients who may have IEIs by primary care providers is paramount to reduce the risk of irrevocable sequelae of invasive infections, such as bronchiectasis, encephalitis, or kidney failure. A number of efforts have attempted to codify a set of “warning signs’’ that offer guidance to primary care doctors. Most recognizable are the “10 Warning Signs’’ that have been widely disseminated by the Jeffrey Modell Foundation for two decades[22]. Before EHRs, the broad phenotyping necessary to assemble a proper picture of heterogeneous IEIs like CVID was not possible, and so guidelines had to be developed by committees and expert opinions. These warning signs largely emphasized infections as a core feature of IEIs. Our results suggest that phenotypes of inflammation, autoimmunity, malignancy, and atopy should also be included. Indeed, two analyses found these 10 warning signs were unable to identify many subjects with known IEIs[23,24], possibly because phenotypes aside from infections were missing. When the warning signs were applied to adults versus children with known IEIs, adult patients were often missed (45% sensitivity for adults versus 64% for children)[25], suggesting the need to modify assessments based on age. In other studies, the need for intravenous antibiotics, failure to thrive, or a relevant family history was found to be the only strong predictors of IEIs[26,27]. An algorithm developed by the Modell Foundation improved IEI diagnoses by using a summation of diagnostic codes[28] and another recent algorithm that summed weighted ICD codes further improved diagnoses[29]; however, these approaches did not include laboratory values. Retrospective gathering of features like we performed here has been useful in aggregating the phenotypic features of patients with IEIs into a score that can discern those with IEIs from those with secondary immunodeficiencies [30]. Recent work used a Bayesian network model to score “risk” in a framework that categorized individuals into either high, medium, or low-risk categories of having any IEI [29]. Their approach also classified each patient into a likely IEI categorization (e.g., combined immunodeficiency, antibody deficiency, etc.). One limitation in Bayesian analyses is in the assessment of probabilities (and conditional probabilities) for rare events; this concern was partially alleviated by employing a large cohort of children with known IEIs. But as a result, that work suggested that 1% of all patients were at medium-to-high risk of IEI, potentially overestimating the true prevalence by 100-200 fold. That work highlights one of our limitations, too, that ascertaining a proper threshold for risk scores is fraught. Regardless, these efforts showcase both the potential and the unmet need for identifying previously undiagnosed patients in large healthcare systems.

There are several inherent limitations to our study. The prediction algorithm is derived primarily from ICD codes within the EHR. Although ICD codes represent an international standard, the specific patterns of assigning ICD codes can often vary across physicians and institutions[31,32]. We overcame this concern by employing phecodes, a generalization of phenotypes derived from ICD codes and better suited for EHR research [33]. However, even using phecodes requires a careful examination of their level of descriptive granularity. For example, one clinical description for CVID in OMIM includes hypothyroidism as a potential phenotypic feature. Accordingly, we utilized the phecode for “Hypothyroidism” (phecode 244.2) in the prediction model. However, no individuals within the CVID cohort at UCLA had this phecode within their medical records. Upon further inspection, we found that this symptom was instead attached as the phecode under “Hypothyroidism NOS” (phecode 244.4). The lesson was that many phecodes under 244.X could equally apply, and that small deviations in diagnosis coding practices could have a large impact on algorithmic outputs. We ameliorated these deviations by not only utilizing symptoms provided in OMIM, but also by learning important model features directly from the training data.

Another limitation of our work was the amount of longitudinal information available in the EHR. Patients move frequently and obtain care from a variety of settings (private practices, urgent care clinics, in addition to large health systems); consequently, only a subset of their data are contained in the health system’s database. Because EHR vendors change with regular occurrence, many EHRs hold only a maximum of 5-15 years worth of data, which may not be enough to fully glean the necessary details of a patient’s health trajectory. We also did not consider the number of times a specific diagnosis appeared nor the order that the phecodes appeared on the medical record. Since CVID is characterized by recurrent infections, we believe that longitudinal information of multiple occurrences would increase the specificity of the model by disregarding individuals with single acute diagnoses. We also did not restrict the types of encounters when collecting the diagnosis codes (e.g., hospitalization, emergency department, or outpatient clinic). Annotations of past and present ICD codes vary considerably across these settings. Instead, we wanted to use as much information as possible to increase the power of our model. However, limiting diagnoses that occur specifically during appointments or hospital visits (as opposed to laboratory tests) could also increase specificity and better differentiate individuals with other immunocompromising conditions (e.g. cancer). We hope to develop these extensions as future work.

Genetic sequencing in patients with immunodeficiency can alter disease management, treatments, and clinical diagnosis [34]. Our approach can be used to expedite the referral of patients to immunologists and to support the need for genome sequencing. By broadening the base of patients studied and their phenotypes, such efforts should expand our understanding of the immunogenetic basis of antibody deficiencies like CVID. In the future, we want to investigate the variants associated with the highest risk scores. The impact of our work will greatly benefit the IEI community as there is an urgent need for more systematic, resource-efficient ways to identify and categorize patients with IEIs.

## Methods

### Study population and electronic health record (EHR) data

The data for this study was extracted from the Discovery Data Repository (DDR) at the University of California, Los Angeles (UCLA). This data warehouse contains all UCLA Health patient information since the implementation of the EHR system in March 2013. The data includes various measurements and metrics such as laboratory tests, medications, billing codes, admissions, and others.

To assess the generalizability of using PheNet scores for different facilities that did not participate in model training, we conducted validation on the UC Health Data Warehouse. The clinical data of the five University of California medical centers contain EHR for 4.97 million patients. We computed the PheNet score for all of them. For computing PheNet scores, we excluded the ICD codes for CVID (‘279.06’, ‘D83.9’, ‘Z94.2’, and ‘Z94.3’) for consistency during determining the phenotypes of patients.

### CVID case definition

Central to our approach was establishing a collection of patients with a known CVID status that served as our “ground truth” cohort. These CVID cases were selected by manual chart review through the following process. First, a chart review of patients with the ICD-10 code D80.* (Immunodeficiency with predominantly antibody defects) helped broaden our ability to study immune disorders under an IRB-approved protocol. This search accumulated 3,200 individuals who all fell under the category of “certain disorders involving the immune mechanism”. Medical records were reviewed to determine the significance of those individuals with this recurring diagnosis code. This process helped eliminate patients who received an immunodeficiency code based on acute occurrences of low antibodies or for access to immunomodulatory treatments including Immunoglobulin (IVIG) without a phenotype of an immune disorder. Additionally, many patients with a cancer diagnosis were excluded based on immunosuppressive medications causing immune dysregulation. The resulting list consisted of 197 patients with CVID who can be consented to research (Supplementary Figure S1).

### Control cohort construction

We constructed a case-matched (see “CVID case definition”) control cohort using the following procedure. Out of the possible N=880K patients in the UCLA Health EHR, we selected individuals based on the self-identified sex, self-identified race/ethnicity, age (closest within a 5-year window), and the number of days recorded in the EHR (closest within a 180-day window) that matched each individual in the case cohort. For age, we used the age listed on the individuals’ most recent encounter. The resulting procedure resulted in a total of N=197 cases and N=1,106 controls.

### Mapping CVID clinical definition to phecodes

To represent features derived from the EHR in our model, we encoded features as phecodes using the ICD code to phecode mapping v1.2[18,35]. These codings represent groupings of ICD codes developed to better represent phenotypic and clinical significance from the EHR and were originally used for phenome-wide association studies. To systematically select the set of phecodes describing CVID, we utilized the entries for CVID listed in the Online Mendelian Inheritance in Man (OMIM) catalog [19] which provides clinical descriptions for thousands of rare diseases. Specifically, we selected the following OMIM ids: 607594 (CVID1), 616576 (CVID12), 614700 (CVID8), 240500 (CVID2), 615577 (CVID10), 616873 (CVID13), 613495 (CVID5), 613494 (CVID4), 617765 (CVID14), 613493 (CVID3), 613496 (CVID6), 614699 (CVID7), 615767 (CVID11). We then used a previously defined database annotating syndromes listed in OMIM with Human Phenotype Ontology (HPO) terms, a set of terms used to clinically describe human phenotypic abnormalities [36,37]. Using this database, we were able to systematically aggregate a list of HPO terms for CVID derived from the clinical descriptions within OMIM. We then used a previously defined mapping between HPO terms and phecodes [12] to translate the list of HPO terms into a list of phecodes which could be constructed using information directly from the EHR. Altogether, this process resulted in a total of 34 unique phecodes describing CVID.

### Selecting model features derived from training cohorts

In addition to using features derived from OMIM (see “Mapping CVID clinical definition to phecodes”), we also include features learned specifically from the training cohort. Although features derived from OMIM may broadly categorize the disease, leveraging information specific to the training cohort can add additional information not already encoded within OMIM. For example, there is variation in how institutions encode diagnoses within the EHR which may not be captured in all OMIM clinical descriptions. Additionally, OMIM definitions are often derived from a limited number of cases due to the rare nature of the diseases. Thus, some symptoms listed might only actually appear in a small percentage of cases since the definitions were derived from such a limited sample size. Other symptoms not currently listed in the clinical descriptions could also be indicative of the disease, but again were not formally added to the clinical definition because the symptoms did not appear in the original samples used for the OMIM description.

To select cohort-specific features, we considered all phecodes present on the medical records of individuals in the training cohort. From a possible 1,800 phecodes, we limited our selection to phecodes present in at least 2 CVID cases and excluded phecodes already selected from OMIM. We then selected the most highly enriched phecodes within the training cohort. We performed a hypothesis test testing the difference in proportions between the case and control groups for each phecode. Ranking phecodes by *p*-value, we selected the top K phecodes. In practice, we set K=10 but also explore alternative values (Supplementary Figure S2A).

### IgG laboratory tests

The final feature included in the model is measurements of Immunoglobulin G (IgG) levels, a common type of antibody. Low IgG is a characteristic of immunocompromised individuals with diseases such as CVID. Instead of using the raw measurement as a feature directly, we convert the values to a categorical scale where the lab value is encoded as ‘0’ if the individual has never received an IgG test, ‘1’ if the individual has had an IgG test >=600 mg/dL (normal range), and ‘2’ if the individual has had an IgG test <600 mg/dL (abnormal). If an individual had multiple recorded IgG tests, we selected the lowest recorded value.

### Model inference

For benchmarking experiments, we performed 5-fold cross-validation within each experiment to quantify the accuracy of various inference frameworks. To address the imbalance of cases in our dataset, we created a more balanced training dataset using random upsampling with an upsampling ratio of 0.50 and downsampling controls to N=10,000. We explored the tradeoff of various upsample ratios and downsampling sample sizes in Supplementary Figure S3, S4. We estimated the weight of each feature using logistic regression (no penalty). We performed additional experiments to quantify performance using a variety of other inference methods, such as ridge regression, random forest, and the inverse-log frequency weighting scheme employed by PheRS (Supplementary Figure S3, S4). Hyperparameters utilized in the ridge regression and random forest models were selected using an additional 5-fold cross-validation step within the training step.

### Comparison with previous methods

We compare PheNet with the current state-of-the-art method PheRS[12] and a related phenotype-risk score that identifies patients who would benefit from chromosomal microarray testing (CMA-score)[13]. Both methods also utilize phecodes as features. PheRS selects phecodes that correspond to the OMIM clinical description of a given disease and then computes the log-inverse frequency of the phecode measured in the general patient population. This is then used as the feature weight in the algorithm and the prediction score is a weighted sum of the weights and the presence of a given phecode, making this approach an entirely unsupervised method that does not leverage any labeled case information. To compare methods, we used PheRS weights computed using the UCLA EHR from over N=880K patients. The CMA-score framework utilizes counts of all possible phecodes and a random forest model. This method requires a training dataset composed of individuals with confirmed CMA tests which was not available to us at this time. Instead, for computing the CMA scores, we utilized pre-computed weights from the original CMA-score study conducted at Vanderbilt University. To quantify the performance of PheNet, PheRS, and the CMA-score, we compute the risk scores for all patients using each method with 5-fold cross validation.

### ICD-based diagnosis date

Although all individuals in the case-cohort were verified to have CVID, we were not able to directly obtain the exact date of diagnosis from the manually reviewed records. For those individuals for whom we could not discern an exact date of diagnosis, we used a heuristic to estimate the date of diagnosis based on occurrences of the ICD code for CVID (D83.9). We refer to this as the “ICD-based diagnosis date” to clarify that it does not constitute the precise date of a formal clinical diagnosis. However, there were 8 individuals who did not have an ICD code for CVID and thus we could not provide an estimated diagnosis date.

### Assessing PheNet using retrospective HER

We first encoded a patient’s most recent visit as time point 0. We recorded the time of an encounter in a patient’s medical record as the number of days before their most recent visit. This provided us a common metric of time to use when performing analyses across all patients. We computed a patient’s PheNet score at 30-day intervals, spanning approximately 6 years (30 days x 12 months x 6 years). At each interval, we only considered features that were recorded up to and including that time point (and time before the given interval). To compute the score percentile for each CVID patient, we used the scores of all other patients taken at time point 0 (i.e. most recent visit) and then added the score of the single CVID patient from the designated time point. Using this distribution, we computed the score percentile for the specific CVID patient at that time point. This is then repeated for all CVID patients across all time points. Because we only had EHR from 2013, we do this to ensure that the overall distribution of scores at earlier time points is not skewed since there are many patients that do not have medical records in the electronic system at earlier time points. We then check to see if any CVID patients reached the top of the score distribution at any time point before an individual’s ICD-based diagnosis (see “ICD-based diagnosis date”).

### Clinical validation of individuals identified by PheNet

To validate our approach, we performed a clinical chart review for the top set of individuals prioritized by PheNet. First, we removed all individuals that were deceased or who had phecodes corresponding to solid organ transplants, cystic fibrosis, or human immunodeficiency virus, resulting in the removal of 42,346 individuals. These specific disorders could lead to immunodeficiency and have a similar profile to CVID, but the cause of their immunodeficiency is already explained. We then selected the top 100 individuals identified by PheNet and a control group of 100 randomly selected individuals from the patient population. For external validation in the UC Health data warehouse, we computed the PheNet score for all patients and counted those with ICD codes for CVID (‘279.06’, ‘D83.9’, ‘Z94.2’, and ‘Z94.3’) for consistency.

Clinical charts were directly reviewed by a clinical immunologist in a blinded review who had access to each individual’s full medical record. The two lists were merged and scrambled, and the clinician was not aware of how the list of individuals was generated. Each individual was ranked according to an ordinal scale from 1-to 5 quantifying the likeliness of having CVID where 1 was defined as “near certainty not CVID” and 5 was “definitive as CVID” meaning that the individual met the criteria of a physician diagnosis.

## Data Availability

Individual-level patient records are not available due to privacy limitations. All summary statistics from the study are contained in the manuscript.

## Acknowledgements

We appreciate the help of Lisa Dahm, PhD, Director, UC Health Data Warehouse. We acknowledge funding from the National Institutes of Health / National Institutes of Allergy and Infectious Diseases (R01 AI153827).

