## Supplementary Materials for "Electronic health record signatures identify undiagnosed patients with Common Variable Immunodeficiency Disease"

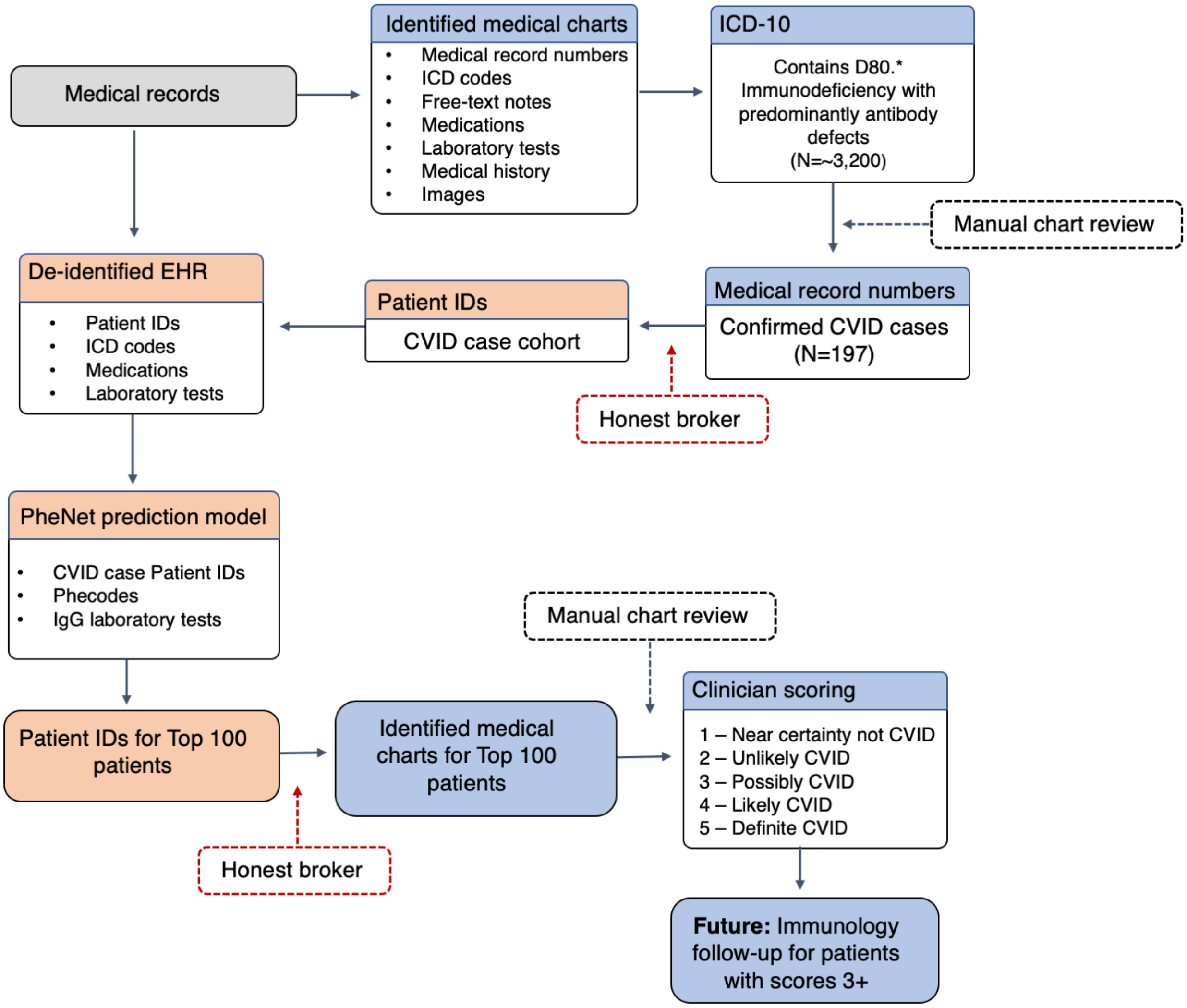


**Supplementary Figure S1: Overview of CVID cohort curation and new CVID patient identification.** We provide a flowchart describing the EHR review process for constructing a well-curated list of clinically diagnosed patients with CVID. We then demonstrate how this cohort is used for training a prediction model which is then used to identify undiagnosed CVID patients in a discovery cohort. A manual chart review is performed on the patients with the highest risk score with the future goal of highly probable CVID patients being referred to an immunologist.

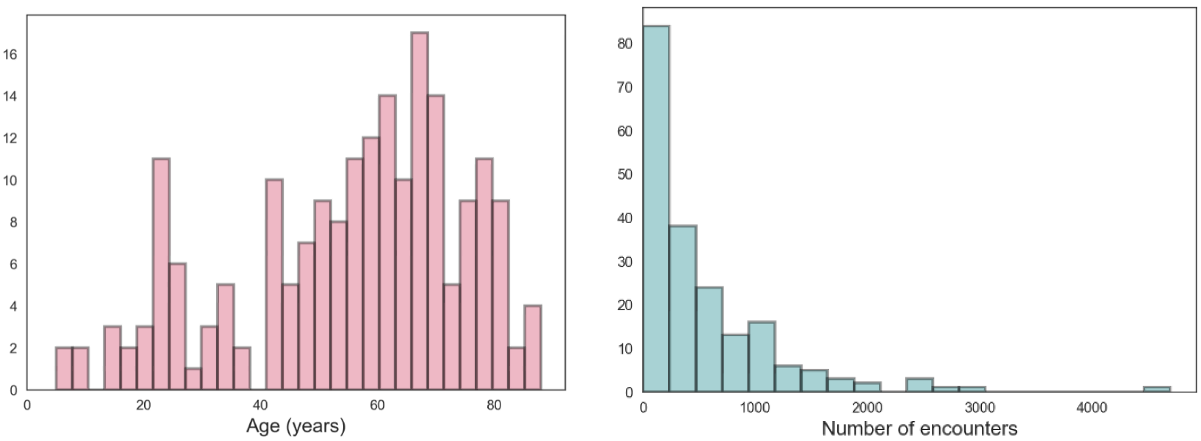


**Supplementary Figure S2: Overview of CVID cohort.** In (A), we show the distribution of ages in the CVID case cohort (N=197). We show the age of patients from their most recent encounter (up to 2019). (B) shows the distribution of the number of encounters recorded in the EHR within the cohort.

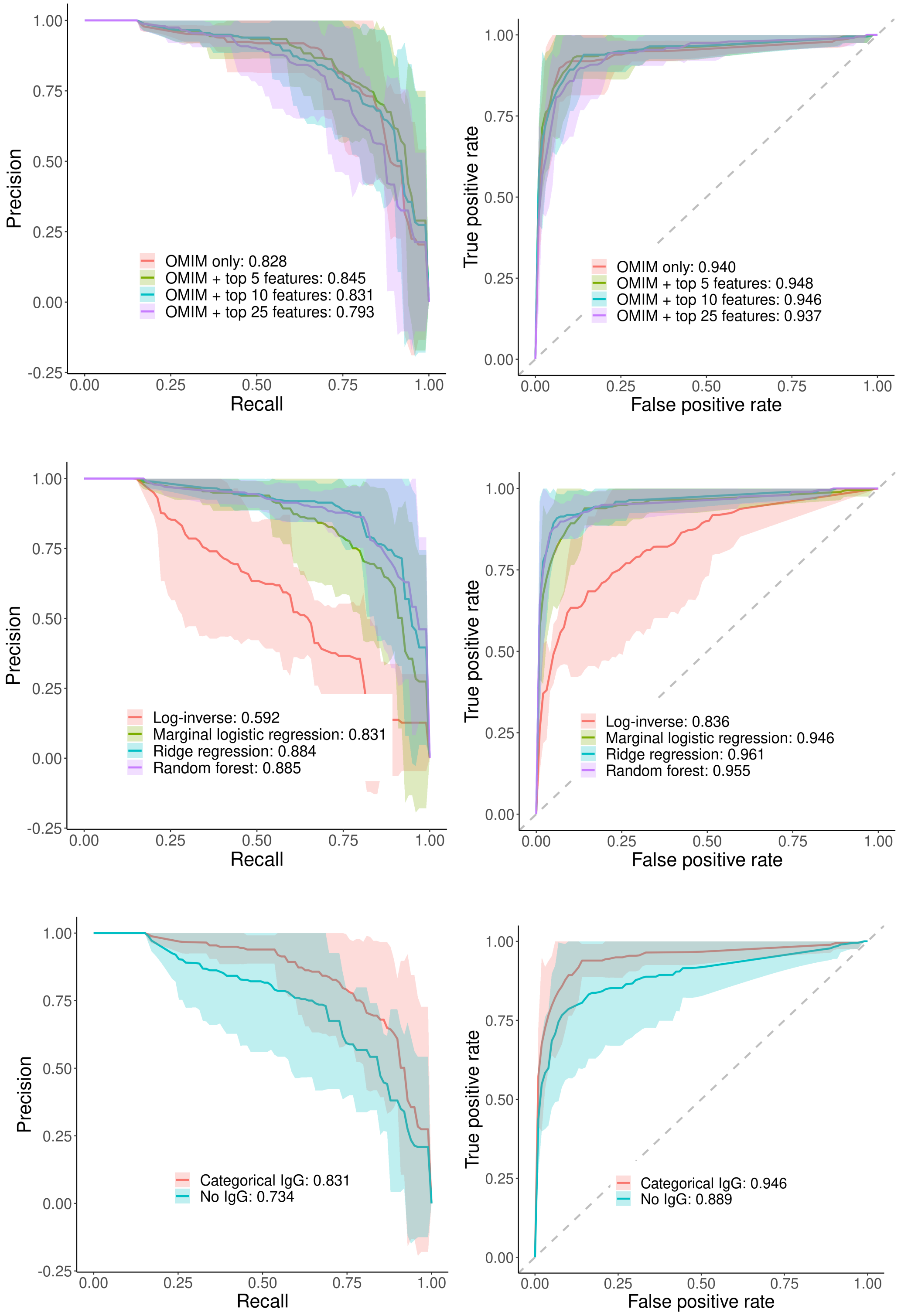

Precision
Recall
OMIM only: 0.828
OMIM + top 5 features: 0.845
OMIM + top 10 features: 0.831
OMIM + top 25 features: 0.793
True positive rate
False positive rate
OMIM only: 0.940
OMIM + top 5 features: 0.948
OMIM + top 10 features: 0.946
OMIM + top 25 features: 0.937
Precision
Recall
Log-inverse: 0.592
Marginal logistic regression: 0.831
Ridge regression: 0.884
Random forest: 0.885
True positive rate
False positive rate
Log-inverse: 0.836
Marginal logistic regression: 0.946
Ridge regression: 0.961
Random forest: 0.955
Precision
Recall
Categorical IgG: 0.831
No IgG: 0.734
True positive rate
False positive rate
Categorical IgG: 0.946
No IgG: 0.889

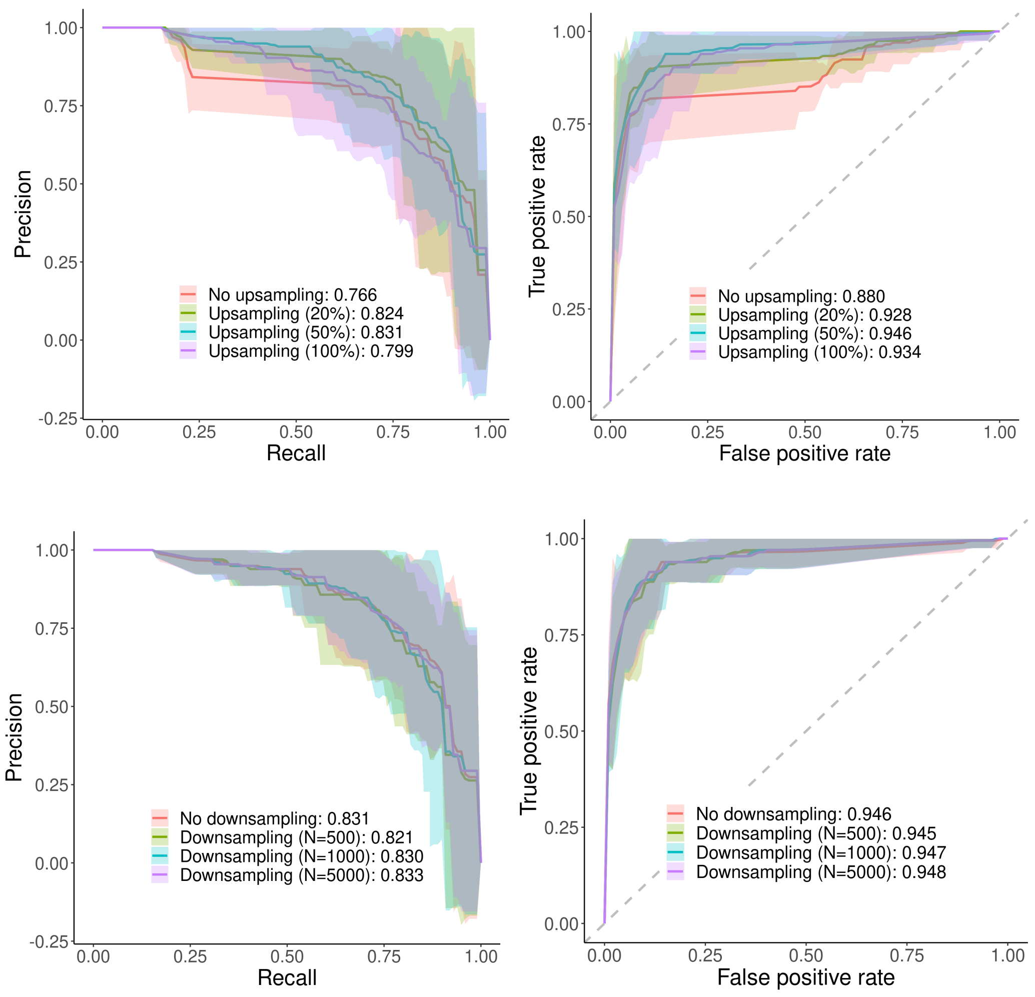


**Supplementary Figure S3: Exploration of model parameters for PheNet**. We show AUC-ROC and AUC-PR curves for the PheNet model using a matched case (N=197) and control cohort (N=1,106) with 5-fold cross-validation. We vary the (A) number of additional phecode features in addition to OMIM-selected features, (B) prediction model, (C) inclusion of immunoglobulin G (IgG) tests, (D) upsampling, and (E) downsampling.

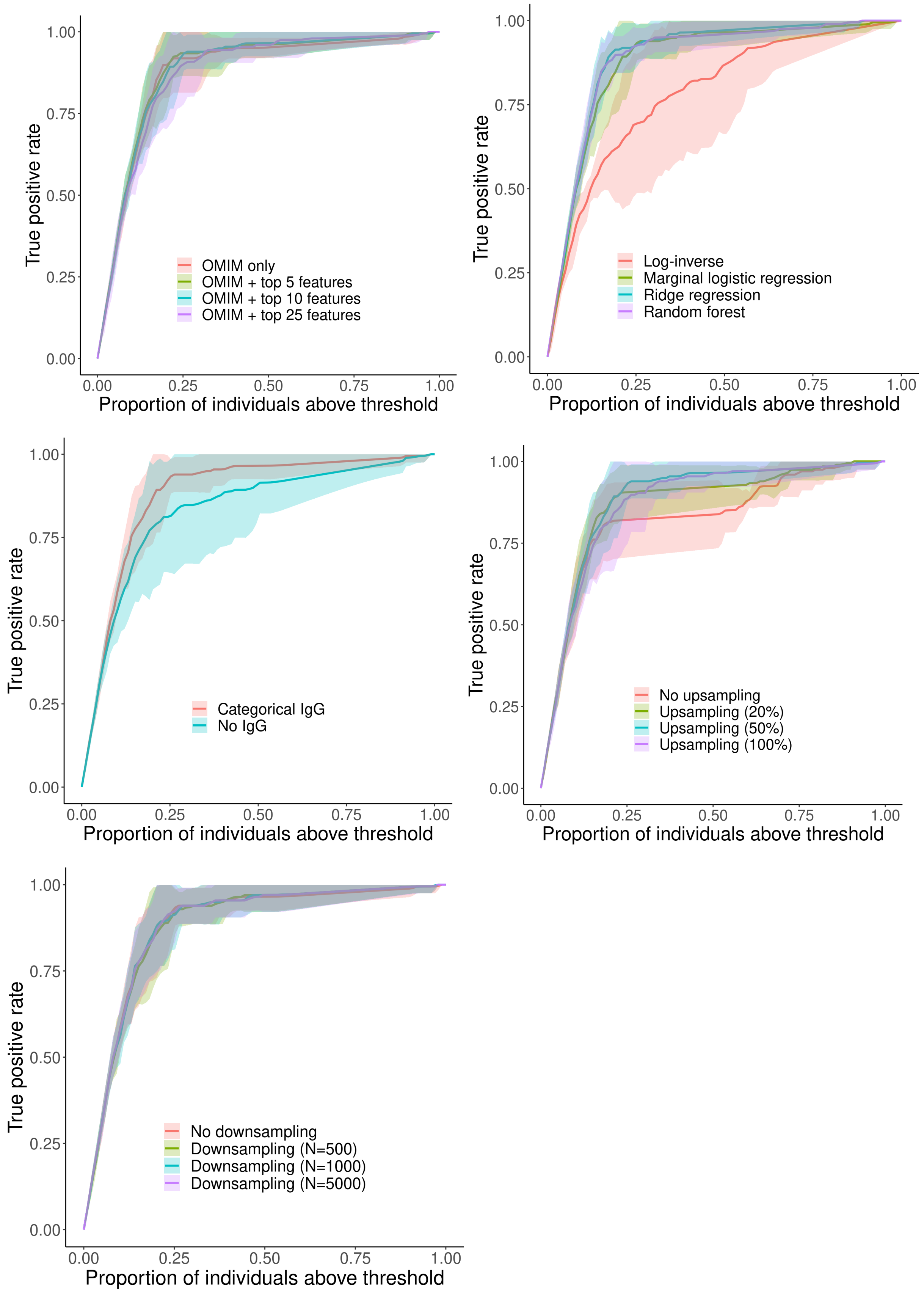
True positive rate
Proportion of individuals above threshold
0.00
0.25
0.50
0.75
1.00
OMIM only
OMIM + top 5 features
OMIM + top 10 features
OMIM + top 25 features
Log-inverse
Marginal logistic regression
Ridge regression
Random forest
Categorical IgG
No IgG
No upsampling
Upsampling (20%)
Upsampling (50%)
Upsampling (100%)
No downsampling
Downsampling (N=500)
Downsampling (N=1000)
Downsampling (N=5000)

**Supplementary Figure S4: Exploration of model parameters for PheNet**. We display a curve showing the proportion of CVID cases captured within the varying percentiles of PheNet scores using a matched case (N=197) and control cohort (N=1,106) with 5-fold cross-validation. We vary the (A) number of phecode features in addition to OMIM-selected features, (B) prediction model, (C) inclusion of immunoglobulin G (IgG) tests, (D) upsampling, and (E) downsampling.

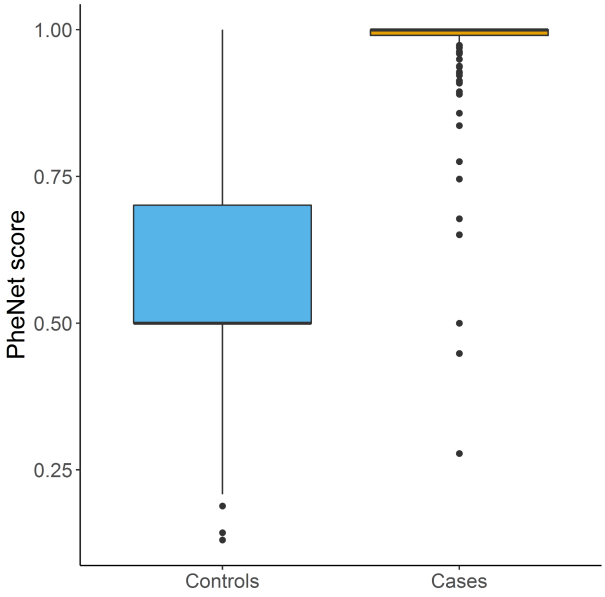


**Supplementary Figure S4: PheNet score distribution between cases and controls.** We show the distribution of PheNet scores within the case-cohort (N=197) and the control-cohort (N=1,106) trained using 5-fold cross-validation. Using a Cochran-Armitage test, we find that the scores in the case-cohort are significantly higher than those in the control-cohort ($p$-value < 2.2e-16).

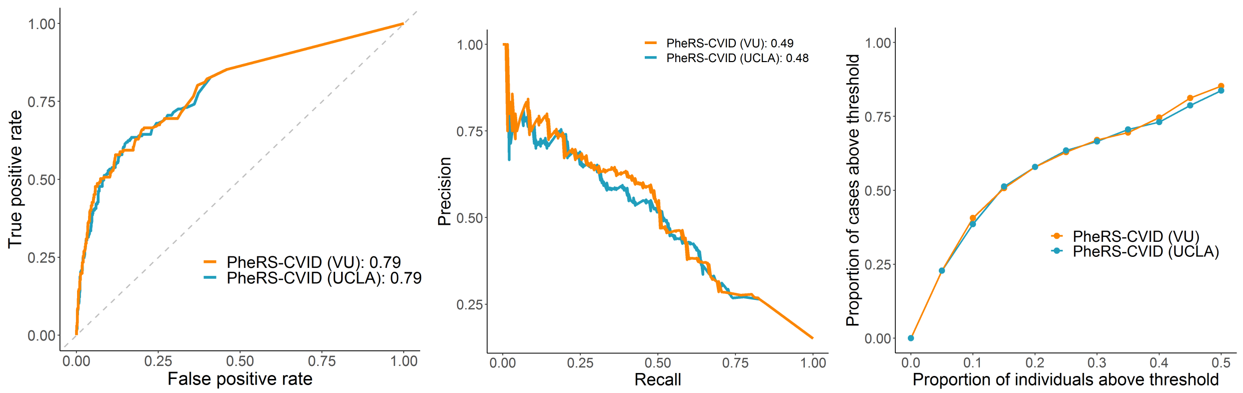


**Supplementary Figure S5: Comparing PheRS performance using models trained at UCLA and Vanderbilt.** We show AUC-ROC, AUC-PR, and calibration curves for the PheRS models trained at UCLA and Vanderbilt (VU). Models were trained and tested using a matched case (N=197) and control (N=1,106) cohort. Because the model is unsupervised, no test-train split was needed.

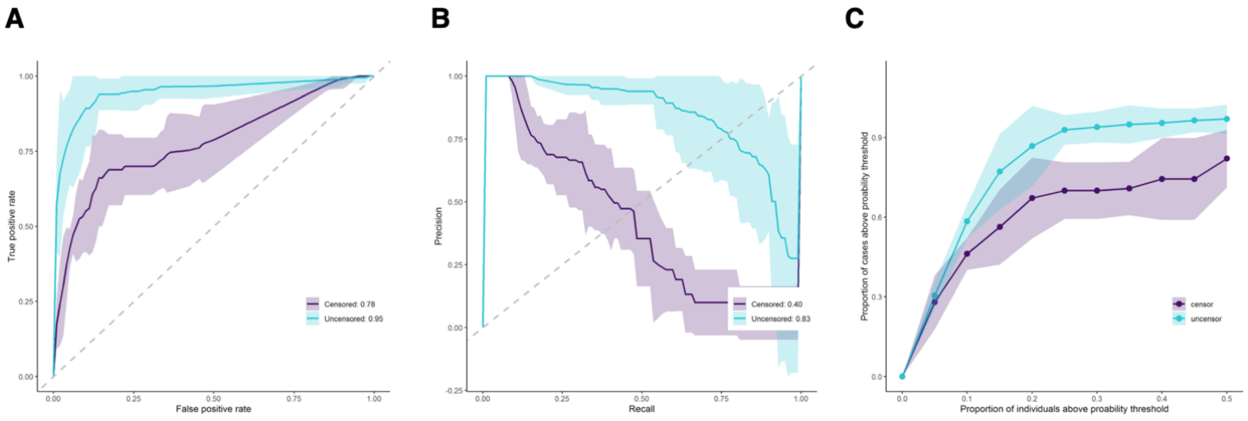


**Supplementary Figure S6: PheNet captures CVID patients with censored data.** Risk scores for CVID were calculated for each patient using PheNet with 5-fold cross-validation across case (N=197) and control (N=1,106) cohorts using both "censored" data that only included information before diagnosis and "uncensored" data that used the entire medical record regardless of diagnosis date. (A) and (B) show AUC-ROC and AUC-PR curves for each model. In (C), patients were ranked, and we report the percentage of CVID cases captured at varying percentile cutoffs. A scoring threshold of 0.90 was used. Note the x-axis is reported in log-scale. Intervals represent scores computed from each fold of cross-validation.

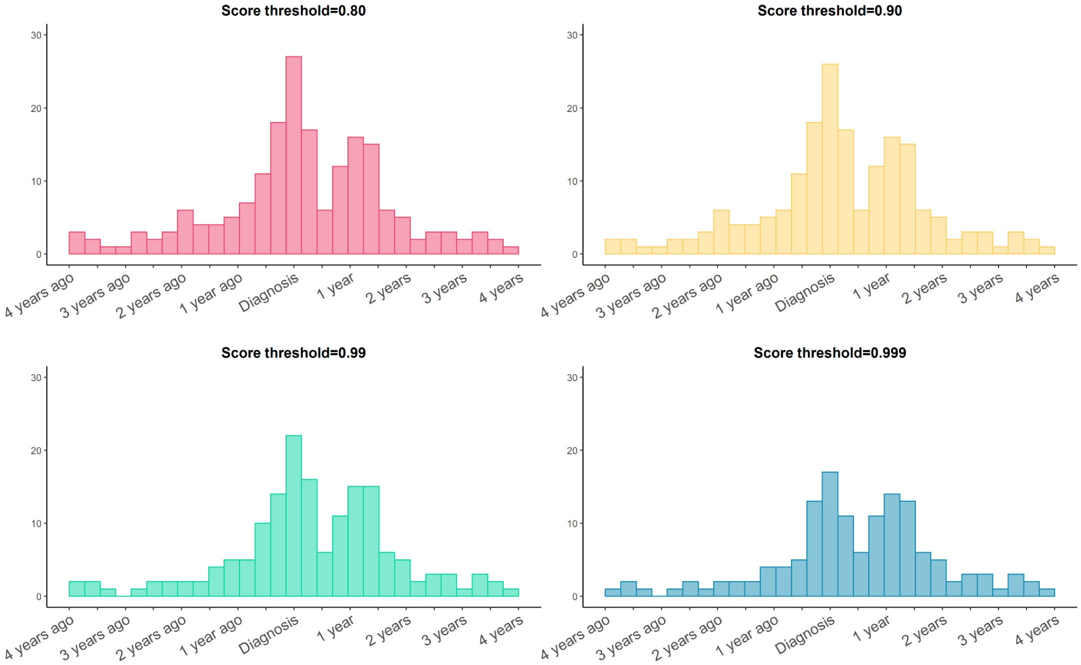


**Supplementary Figure S7: Different score thresholds capture various numbers of CVID patients before diagnosis.** Training on the full UCLA Health population (N=~880K), we estimated PheNet scores for CVID patients using only information before their ICD-based diagnosis and 5-fold cross-validation. We show the distribution of times when patients pass the scoring threshold at 0.80, 0.90, 0.99, and 0.999. We only show CVID patients with at least 1 year of recorded EHR data prior to their diagnosis (N=58).

| | Top 100 | Random 100 |
|---|---|---|
| **Age (years)** | | |
| Mean (s.d.) | 57.35 (20.63) | 43.35 (21.18) |
| Median | 61.00 | 42.00 |
| **Sex (%)** | | |
| Male | 72 | 28 |
| Female | 54 | 46 |
| **Number of ICD codes** | | |
| Mean unique (s.d.) | 242.01 (20.63) | 30.81 (21.18) |
| Median | 210.00 | 18.00 |
| **Medical record length** | | |
| Mean years | 15.52 (7.22) | 7.07 (6.87) |
| Median years (s.d.) | 15.50 | 4.90 |

**Supplementary Table S1: Demographics of top 100 patients identified by PheNet and 100 randomly sampled patients.** We show a summary of the top 100 individuals with the highest PheNet score out of the discovery cohort (N=~880K) and a control group of 100 randomly sampled patients from the patient population. We provide summary statistics on patients' age, self-reported sex, number of unique ICD codes, and the number of years recorded in the EHR.

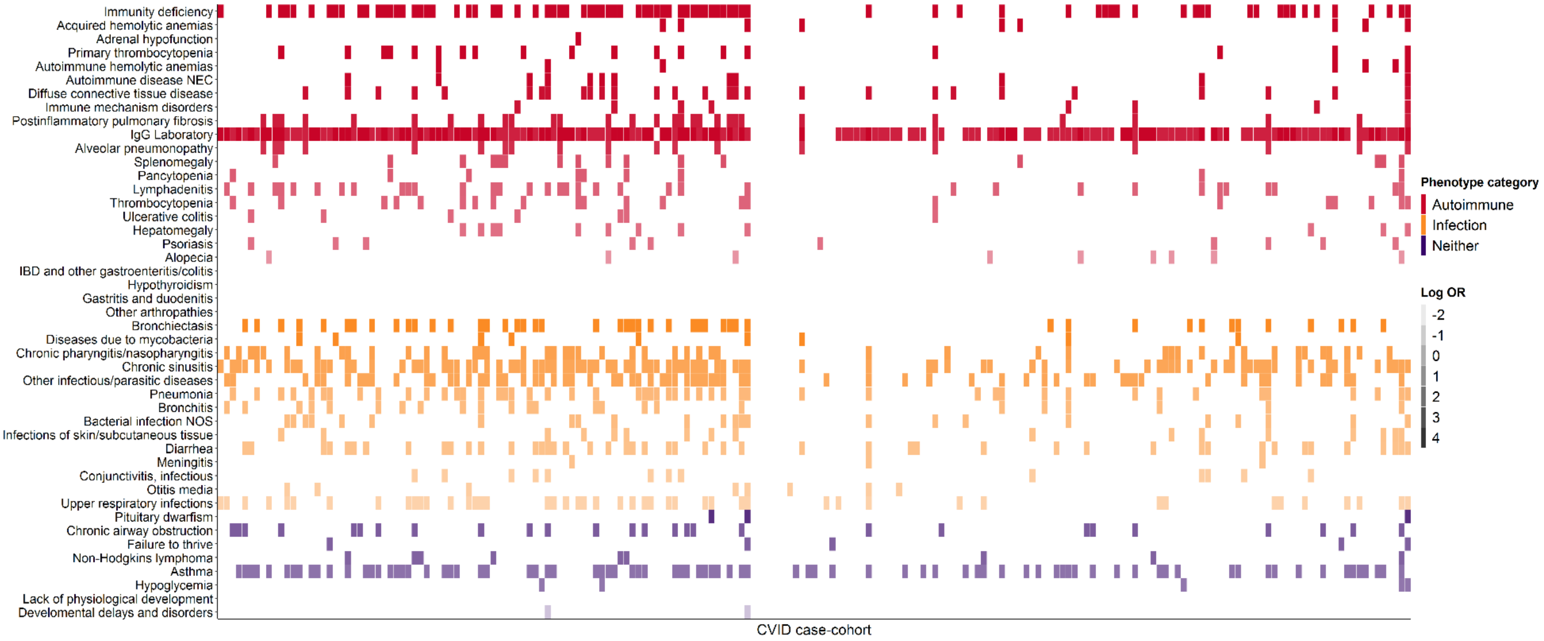


**Supplementary Figure S8: EHR-signatures of individuals in the CVID case cohort.** Each row shows a clinical feature from the PheNet model and each column is a patient's EHR-profile. Individuals in the CVID cohort (N=197) are shown where the lowest to highest risk scores are displayed left to right. Boxes are colored according to phenotype category (autoimmune, infection, neither) and shaded according to the weight of each feature in the algorithm in the form of log odds ratio.

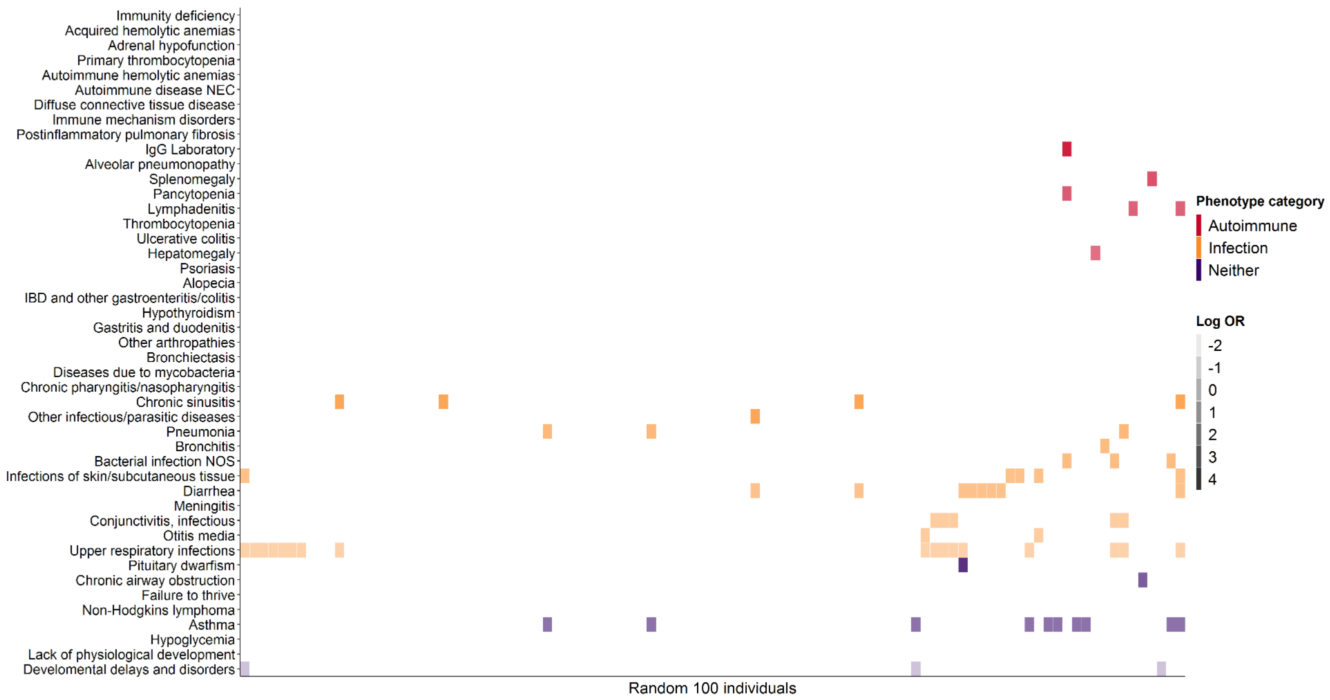


**Supplementary Figure S9: EHR-signatures of individuals in a randomly selected sample.** Each row shows a clinical feature from the PheNet model and each column is a patient's EHR-profile. The 100 individuals randomly sampled from the patient population are shown where the lowest to highest risk scores are displayed left to right. Boxes are colored according to phenotype category (autoimmune, infection, neither) and shaded according to the weight of each feature in the algorithm in the form of log odds ratio.
